# Risk factors and protective factors of mental health during COVID-19 outbreak and lockdown in adult Indian population- A cross-sectional study

**DOI:** 10.1101/2020.06.13.20130153

**Authors:** Jayakumar Saikarthik, Ilango Saraswathi, Thirusangu Siva

## Abstract

**Background:** The novel Corona virus has derailed the entire world and various steps have been taken by the health authorities to tackle this pandemic. Nationwide lockdown has been imposed to control the spread of COVID-19 outbreak in India, which could have psychological impact on the population.

**Aim:** Our study aims to study the effect of the COVID-19 outbreak & subsequent lockdown on mental health status of adult Indian population along with identifying the high-risk groups.

**Methodology:** An online survey was conducted during 3^rd^ phase of lockdown gathering details about sociodemographic variables, practice of precautionary measures, awareness and concerns regarding COVID-19 and mental health status of the participants through DASS21 questionnaire from 873 adults.

**Results:** The prevalence of depression, anxiety and stress were 18.56%, 25.66%, and 21.99% respectively including higher number of participants with mild depression (15.1%) and stress (14.5%) and moderate anxiety (16.3%). Female gender, age <25 years, unemployment, self-business, employed in private sector, lack of formal education, larger household size, parenthood (>2 kids) were associated with increased likelihood of negative mental health. Confidence in physician’s ability to diagnose COVID-19 infection, decreased self-perceived likelihood of contracting COVID-19, lesser frequency of checking for information on COVID-19 and satisfaction of information received were protective against negative mental health.

**Conclusion:** This landmark study identified the protective and risk factors of mental health during COVID-19 pandemic, to help authorities and mental health workers to strategize and deliver interventional methods to maintain psychosocial wellbeing of the population.

## Introduction

COVID-19 outbreak, caused by a novel corona virus, SARS-CoV-2, which originated from China, has spread worldwide, earning the pandemic status by WHO on March 11, 2020 [1]. As of the 1^st^ week of June, 2020, India has emerged as the fifth hardest hit country with 247,000 confirmed cases of COVID-19 and 7000 deaths [2]. Droplets, contact with immediate environment around infected persons including direct or indirect and airborne in specific circumstances are the main proposed routes of transmission [3].

As of now, there are no specific medicines or vaccines available for COVID-19. To tackle this, the Government of India has opted for nationwide lockdown and emphasized on social distancing. Though quarantine and lockdown help containing the spread of infection, it is also accompanied by potential psychological distress in the population. Isolation, fear of contracting the disease, confusion created by rumours, financial strain, apprehension regarding job security, boredom, frustrations, lack of freedom and space due to restrictions, alcohol withdrawal, concerns for the family members that occur during lockdown period could affect the mental health of the population to varying degrees.

Studies conducted during earlier epidemics like SARS, equine influenza, Ebola have noted that there was increased psychological distress due to the epidemic and quarantine [4, 5]. Similarly, studies conducted recently in other countries like China, Italy, Iran have noted increased prevalence of mental health disorders like depression, anxiety, stress and sleep disturbances during COVID-19 outbreak [6-8]. A recent survey conducted on March 2020 in India, has found that more than three fourth of the study participants had self-perceived need for help for their mental well-being [9].

Till date, very few studies have been conducted on the psychological impact of COVID-19 outbreak & lockdown in India, which either focused on specific areas like perceived mental health care need [9] and effect of gender and marital status [10], or conducted on specific population like healthcare workers [11] and pharmacy students [12]. However, there are no studies conducted to assess the impact of COVID-19 outbreak & lockdown on the mental health status of the general population of India with emphasis on the risk factors and protective factors. Our study aims to assess the prevalence of affective components of mental health viz. depression, anxiety and stress along with identifying the high-risk group of population. We believe, our study would help the authorities and mental health professionals in strategizing and delivering mental healthcare to the population targeting on the high-risk group and help maintaining the psychosocial well-being of the Indian population.

## Methodology

### Study design and participants

The study was a cross sectional survey conducted through an online survey platform. The invite link to attend the survey was distributed in social networks like Facebook, WhatsApp and Telegram. Only adult (age above 18 years), Indian residents were invited. The objective of the study was explained, and the consented participants filled out the survey and could quit the survey as and when needed. The entire survey was in English and a tentative average time duration needed to fill out the survey was mentioned beforehand (15-20 minutes). Ethical approval was obtained from Institutional Ethics Committee on Fasttrack basis and the anonymity of the participants was maintained.

### Survey development and Data collection

The survey included a self-administered questionnaire which was developed after extensive literature survey and included questions pertaining to sociodemographic variables and COVID-19 outbreak & lockdown related variables. Snowball sampling method was used, and the data collected between May 5^th^ to 14^th^, 2020 during the third phase of lockdown in India was taken for this study. The data collected were sociodemographic variables, COVID-19 outbreak & lockdown related variables and mental health status of the participants.

#### Sociodemographic variables

Sociodemographic variables included gender, age, educational status, employment status, marital status, monthly income, parental status and household size.

#### COVID-19 outbreak & lockdown related variables

COVID-19 outbreak & lockdown related variables included (a) practice of personal precautionary measures; wearing masks and gloves in public places, frequency of hand washing with soap or sanitizer per day, The participants were asked if they or any of their peers tested for COVID-19 (b) awareness and knowledge regarding COVID-19 pandemic; route of transmission, means of gathering information, frequency of checking for information and level of satisfaction of the attained information, (c) personal concerns regarding the outbreak; level of confidence in the physician to diagnose COVID-19 infection, self-perceived likelihood of contracting COVID-19 and surviving if contracted with COVID-19 and concerns for family members to contract the infection.

#### Psychological status of the participants

Affective component of mental health of the participants viz depression, anxiety and stress were assessed using Lovibond and Lovibond’s short version of the Depression Anxiety Stress Scale 21 (DASS21). DASS21 is a reliable instrument used in clinical and nonclinical samples which can measure and differentiate between the three negative emotional states [13, 14]. The sub-scores for depression, anxiety and stress were summed up and categorized into “normal”, “mild”, “moderate”, “severe” and “extremely severe”. Cut-off score of ≥10 for depression, ≥8 for anxiety and ≥15 for stress were considered to be having the aforesaid disorders [15].

### Statistical analysis

Analysis was performed using SPSS V.26.0, IBM, New York, USA. Descriptive analysis was performed for all variables. Depression, anxiety and stress scores were expressed as mean and SD. Multicollinearity was checked between independent variables and the variance inflation factor (VIF) was found to be less than 3.

To explore potential predictors for depression, anxiety and stress, binomial logistic regression analysis of each independent variable was performed separately, and the results were expressed as crude odds ratio (cOR) with 95% confidence interval (CI) and P value. This was followed by Multivariate binomial logistic regression analysis using ‘stepwise forward LR’ technique, which included independent variables which were found to be significant (P < 0.25, Hosmer-Lemeshow recommendation) by univariate analysis. The regression analysis was performed in two blocks, sociodemographic variables block, and COVID-19 outbreak & lockdown related variables block. The latter block was explored after controlling for the significant sociodemographic factors. The results were expressed as Wald test value, adjusted odds ratio (aOR) with 95% confidence interval and P value (P<0.05 was considered statistically significant).

## Results and Discussion

### Descriptive characteristics of the study population

968 responses were received, out of which 64 responses were incomplete, 25 respondents were underage, and 6 respondents were of a different nationality and were hence excluded. The final sample size was 873. We had an almost even participation from males (54.1%) and females (45.9%) and majority of the participants were of age group 18 to 45 years (85.1%) with the average age of 33.6±12.15 years. The descriptive statistics of the study population is shown in Table 1.

**Table 1:** Descriptive statistics of the study population.

| Variables | Subgroups | Number of participants | % |
| --- | --- | --- | --- |
| **Sociodemographic variables** |  |  |  |
| Gender | Male | 473 | 54.1 |
|  | Female | 400 | 45.9 |
| Age (years) | 18 to 25 | 276 | 31.6 |
|  | 26 to 35 | 225 | 22.7 |
|  | 36 to 45 | 269 | 30.8 |
|  | 46 to 55 | 40 | 4.5 |
|  | 56 & above | 63 | 7.2 |
| Educational status | None | 28 | 3.2 |
|  | Higher secondary school | 261 | 29.8 |
|  | Bachelor's degree | 413 | 47.3 |
|  | Master's degree | 147 | 16.8 |
|  | Doctorate degree | 24 | 2.7 |
| Marital status | Single | 388 | 44.4 |
|  | Married | 469 | 53.7 |
|  | Widowed/ separated | 16 | 1.9 |
| Employment status | Student | 259 | 29.7 |
|  | Employed – Government | 80 | 9.2 |
|  | Employed – private | 309 | 35.4 |
|  | Self-business | 125 | 14.3 |
|  | Unemployed | 100 | 11.4 |
| Monthly income (INR) | 10,000 and less | 307 | 35.1 |
|  | 10,001-20,000 | 42 | 5.4 |
|  | 20,001-30,000 | 31 | 3.5 |
|  | 30,001-40,000 | 75 | 8.5 |
|  | 40,001-50,000 | 59 | 6.7 |
|  | 50,001-100,000 | 207 | 23.3 |
|  | 100,000 & above | 152 | 17.4 |
| Parental status | No kid | 406 | 46.5 |
|  | 1 kid | 141 | 16.1 |
|  | 2 kids | 254 | 29.09 |
|  | 3 or more kids | 72 | 8.2 |
| Household size | 1 member | 12 | 1.3 |
|  | 2 members | 125 | 14.3 |
|  | 3-6 members | 708 | 81.2 |
|  | More than 6 members | 28 | 3.2 |
| **Practice of personal precautionary measures during COVID-19 outbreak and lockdown** |  |  |  |
| How often do you wear | No | 103 | 11.8 |
| mask and gloves while | Sometimes | 109 | 12.4 |
| being outside in public | Yes | 661 | 75.8 |
| places? |  |  |  |
| How often do you wash | less than 5 times | 254 | 29.1 |
| your hands with soap or | 5 to 10 times | 308 | 35.2 |
| hand sanitizer per day? | 10 to 15 times | 227 | 26.1 |
|  | More than 15 times | 84 | 9.6 |
| **Awareness and Knowledge about COVID-19 outbreak** |  |  |  |
| Route of transmission |  |  |  |
| Contact with infected | No | 193 | 22.1 |
| person | Yes | 680 | 77.9 |
| Droplets | No | 252 | 28.9 |
|  | Yes | 621 | 71.1 |
| Airborne | No | 676 | 77.4 |
|  | Yes | 197 | 22.6 |
| Contact with contaminated | No | 282 | 32.3 |
| objects | Yes | 591 | 67.7 |
| Through food and water | No | 816 | 93.5 |
|  | Yes | 57 | 6.5 |
| Pet animals | No | 853 | 97.7 |
|  | Yes | 20 | 2.3 |
| Are you aware of the | Yes | 843 | 96.5 |
| increase in number of COVID-19 cases in India? | No | 30 | 3.5 |

| Source of daily information regarding COVID-19 outbreak |  |  |  |
| --- | --- | --- | --- |
| Internet | No | 227 | 26 |
|  | Yes | 646 | 74 |
| TV | No | 164 | 18.8 |
|  | Yes | 709 | 81.2 |
| Friends and relatives | No | 569 | 65.1 |
|  | Yes | 304 | 34.9 |
| MOH messages | No | 800 | 91.6 |
|  | Yes | 73 | 8.4 |
| Radio | No | 869 | 99.5 |
|  | Yes | 4 | 0.5 |
| Other sources<br>(Newspaper, magazines<br>etc.) | No | 821 | 94 |
|  | Yes | 52 | 6 |
| Are you satisfied with the<br>information received daily<br>regarding COVID-19<br>outbreak? | Highly satisfied | 47 | 5.4 |
|  | Satisfied | 575 | 65.9 |
|  | Not satisfied | 106 | 12.1 |
|  | Highly not satisfied | 34 | 3.9 |
|  | I don't know | 111 | 12.7 |
| How often do you check for<br>information about COVID<br>19 per day? | Less than 5 times | 490 | 56.1 |
|  | Less than 10 times | 205 | 23.5 |
|  | Less than 20 times | 146 | 16.7 |
|  | More than 21 times | 32 | 3.7 |
| Personal concerns regarding COVID-19 outbreak and lockdown |  |  |  |
| How confident are you on<br>your physician to diagnose | Highly confident | 110 | 12.6 |
|  | Confident | 528 | 60.5 |
|  | Not confident | 51 | 5.8 |
| COVID-19 infection? | I don't know | 184 | 21.1 |
| How likely are you to contract COVID-19 during this outbreak? | Highly likely | 16 | 1.8 |
|  | Likely | 248 | 28.4 |
|  | Not likely | 345 | 39.4 |
|  | Highly not likely | 89 | 10.2 |
|  | I don't know | 175 | 20 |
| How likely are you to survive, if contracted with COVID-19 infection? | Highly likely | 129 | 14.8 |
|  | Likely | 456 | 52.2 |
|  | Not likely | 55 | 6.3 |
|  | Highly not likely | 20 | 2.3 |
|  | I don't know | 213 | 24.4 |
| How concerned are you about your family members to contract COVID-19 infection during this outbreak? | Highly concerned | 566 | 64.8 |
|  | Concerned | 276 | 31.6 |
|  | Not concerned | 11 | 1.3 |
|  | Highly not concerned | 12 | 1.4 |
|  | I don't know | 8 | 0.9 |

Average score for the three subscales were, depression: 5.02±4.96, anxiety: 4.41±3.41 and stress: 7.77±7.42. 18.56% had symptoms of depression with the majority having mild depression (15.1%), 25.66% had symptoms of anxiety where, participants with moderate anxiety predominated (16.3%) and 21.99% had symptoms of stress with the maximum in mild stress category (14.5%) (Figure 1). Our results were considerably higher than the prevalence of these negative components of psychological health assessed by studies in the population, before the pandemic [16, 17] while being similar to the findings by studies conducted in other countries during COVID-19 pandemic [6, 8] and SARS pandemic [4].

**Figure 1:**
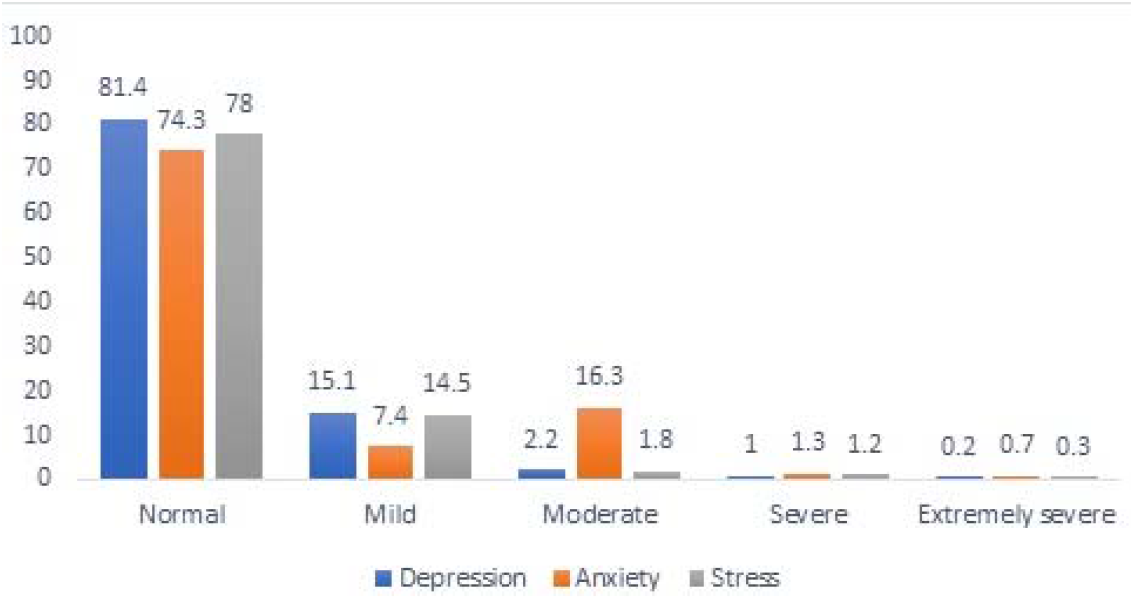
Prevalence of Depression, Anxiety and Stress in the study population (in %)

### Association between sociodemographic, COVID-19 outbreak & lockdown related variables and mental health status

Marital status, wearing masks and gloves in public places, awareness of increase in number of COVID-19 cases, Satisfaction with the information received daily regarding COVID-19 outbreak (Table 2 and 3) were excluded in the final model for depression based on the initial binomial logistic regression.

**Table 2:**
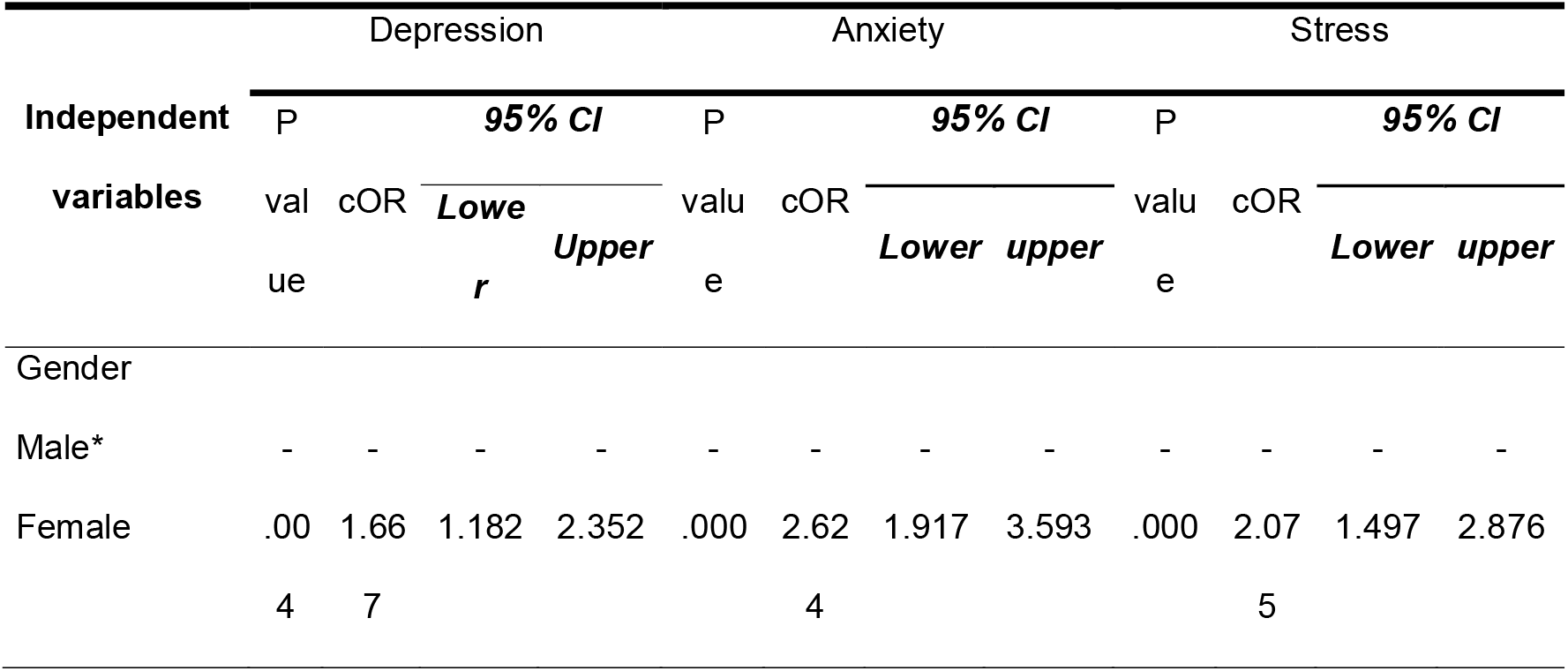
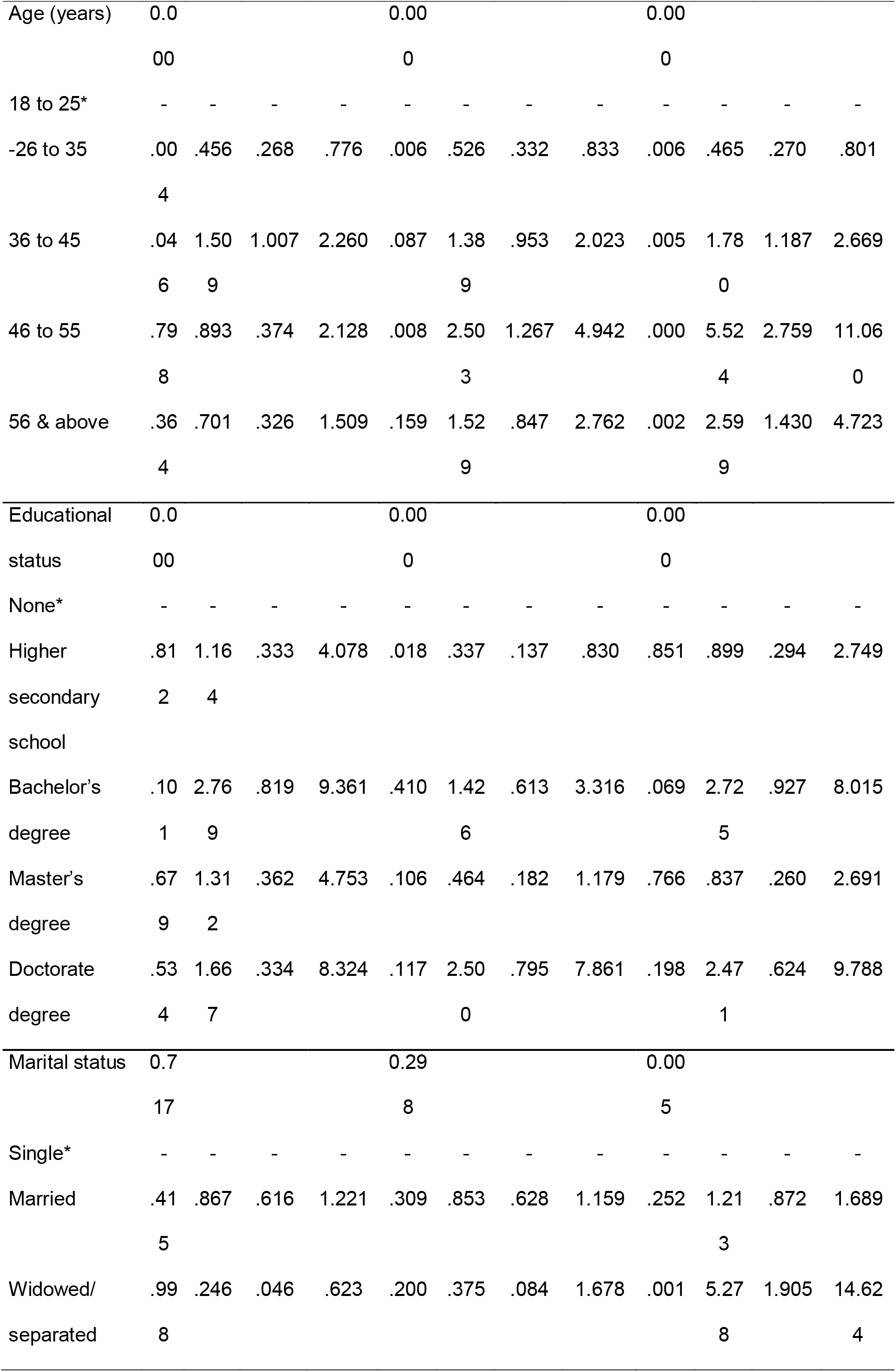
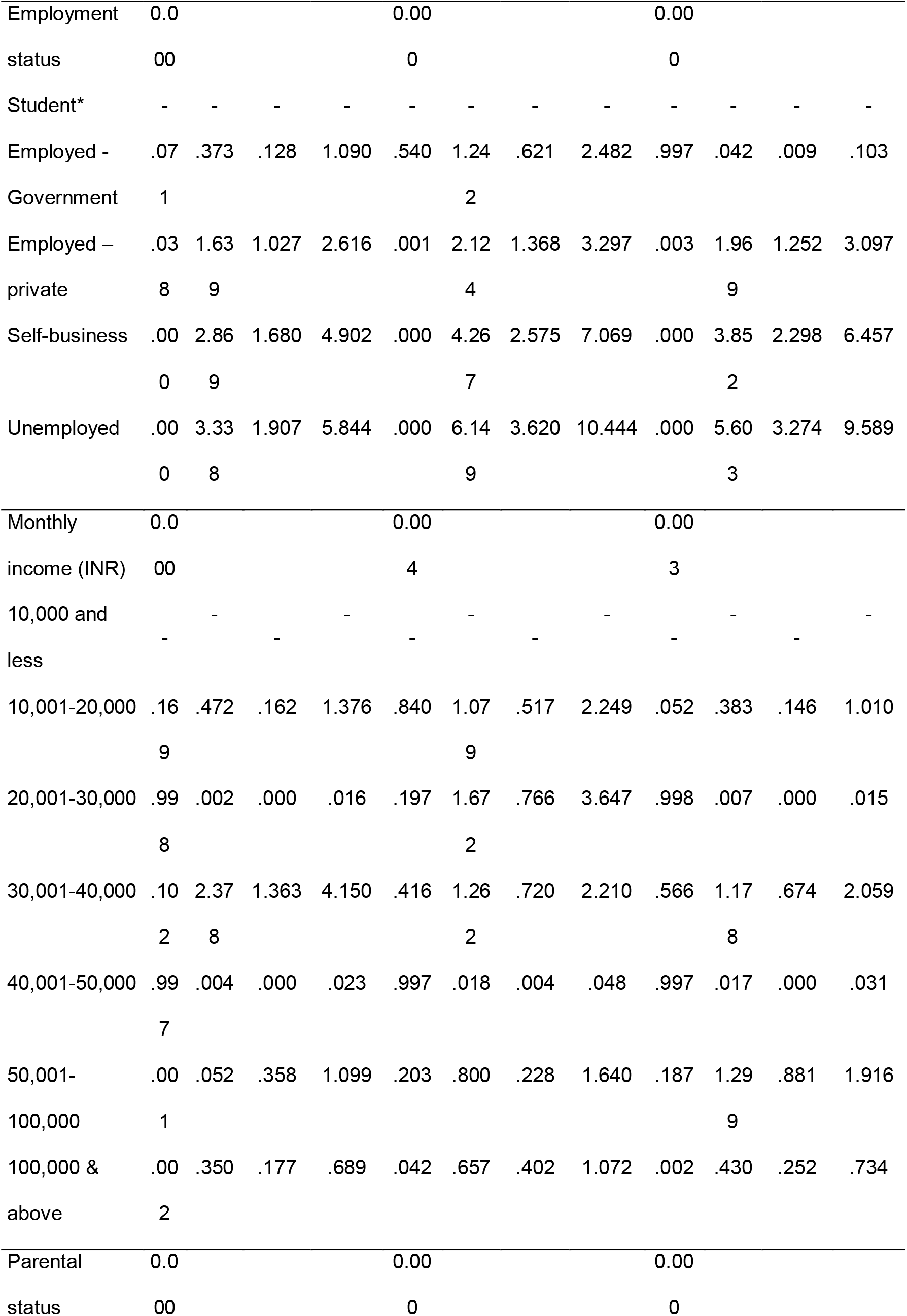
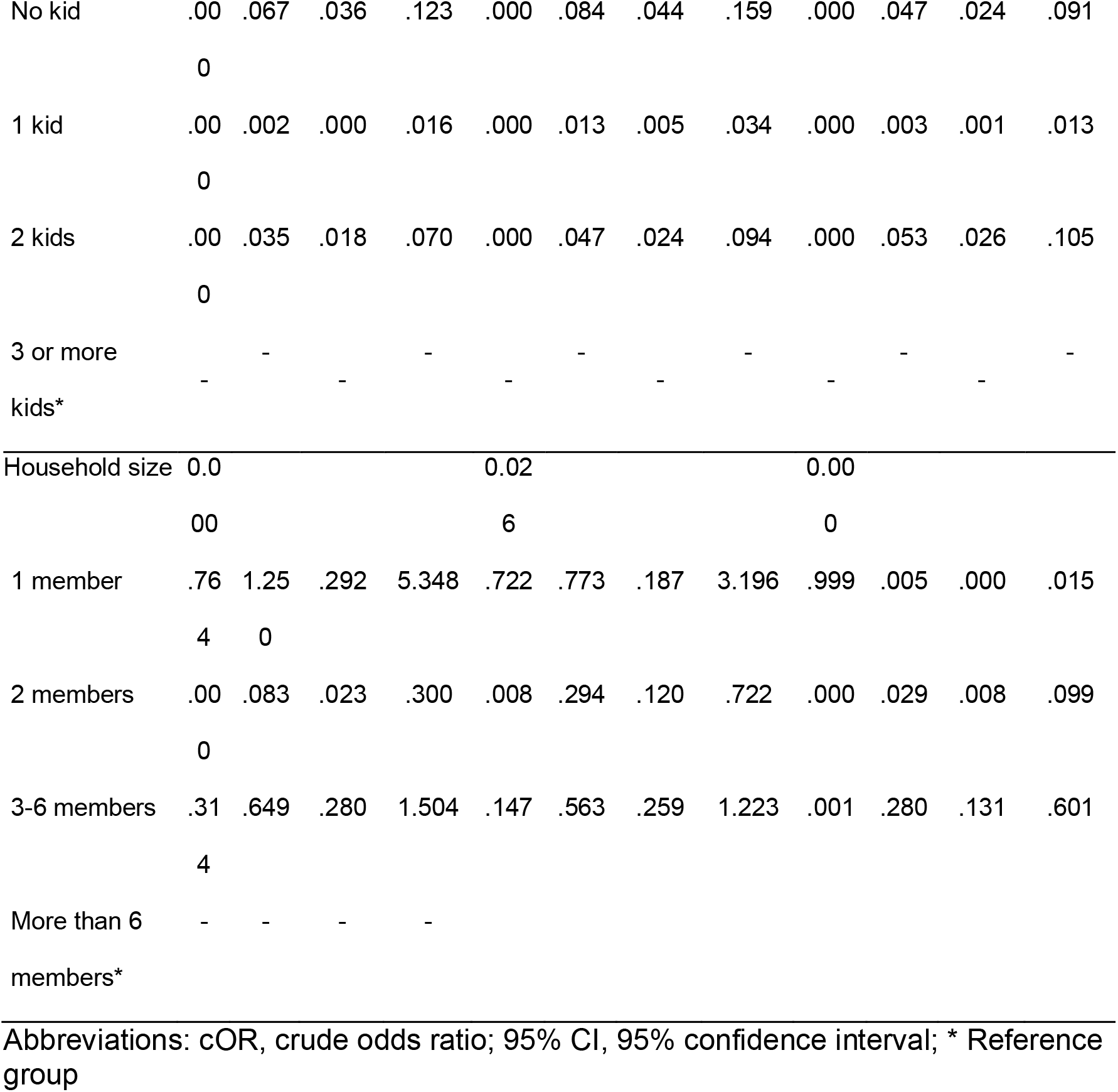
Bivariate analysis of sociodemographic variables by binary logistic regression

**Table 3:**
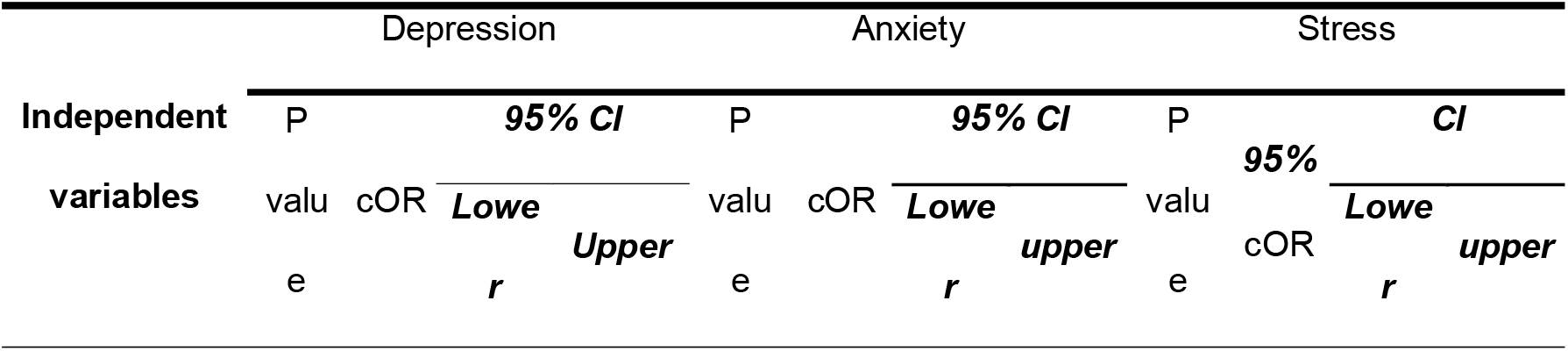
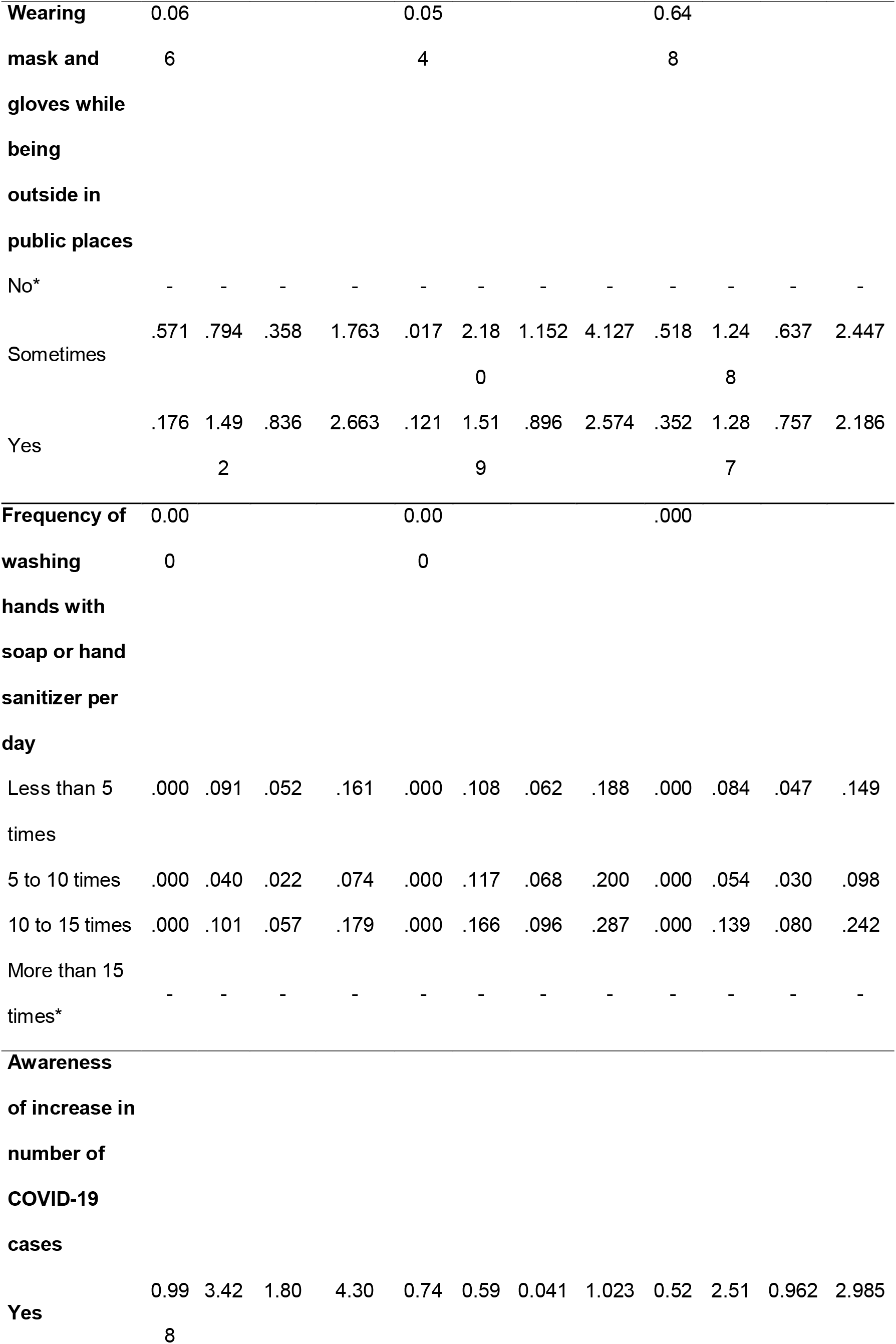
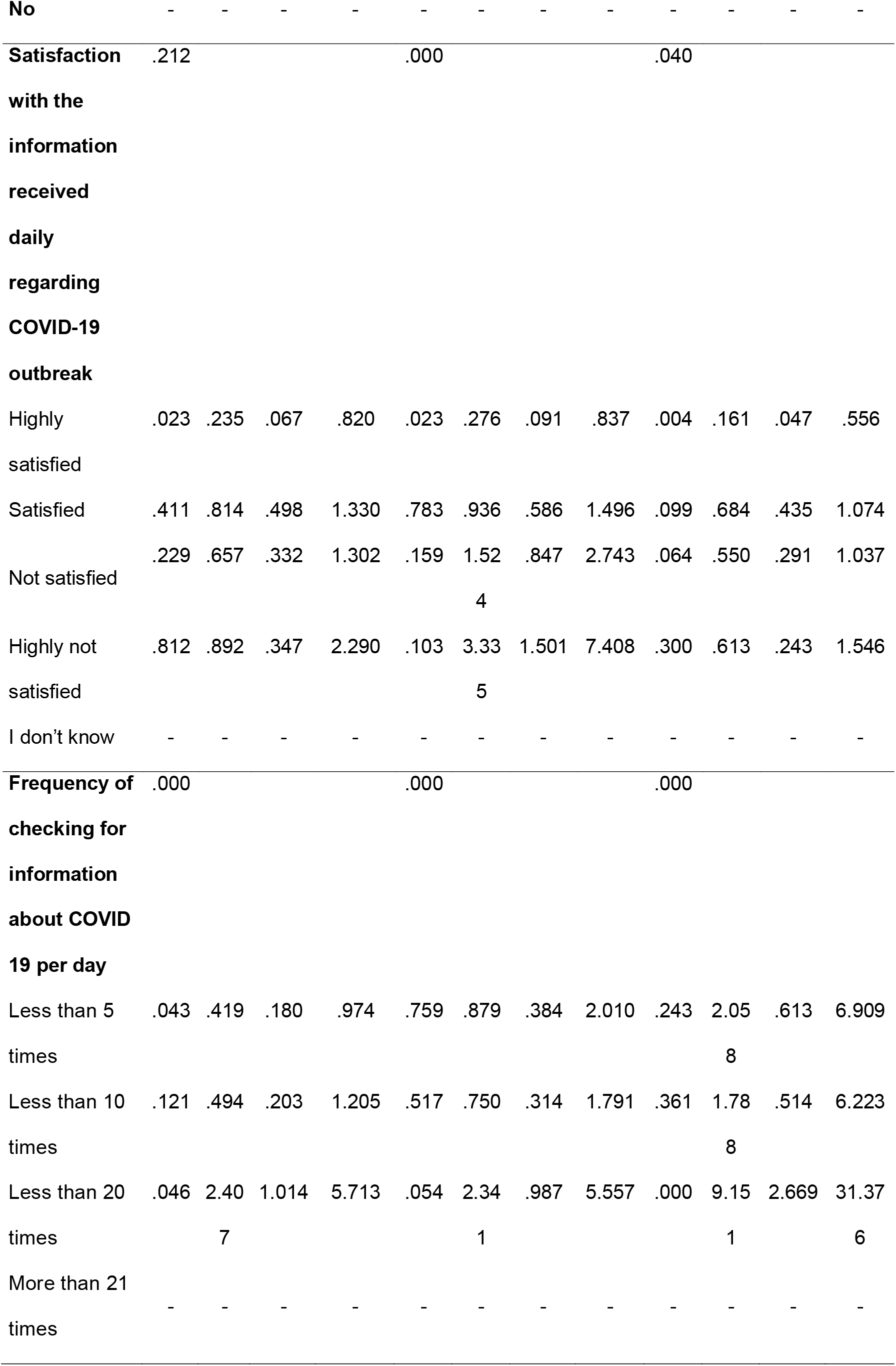
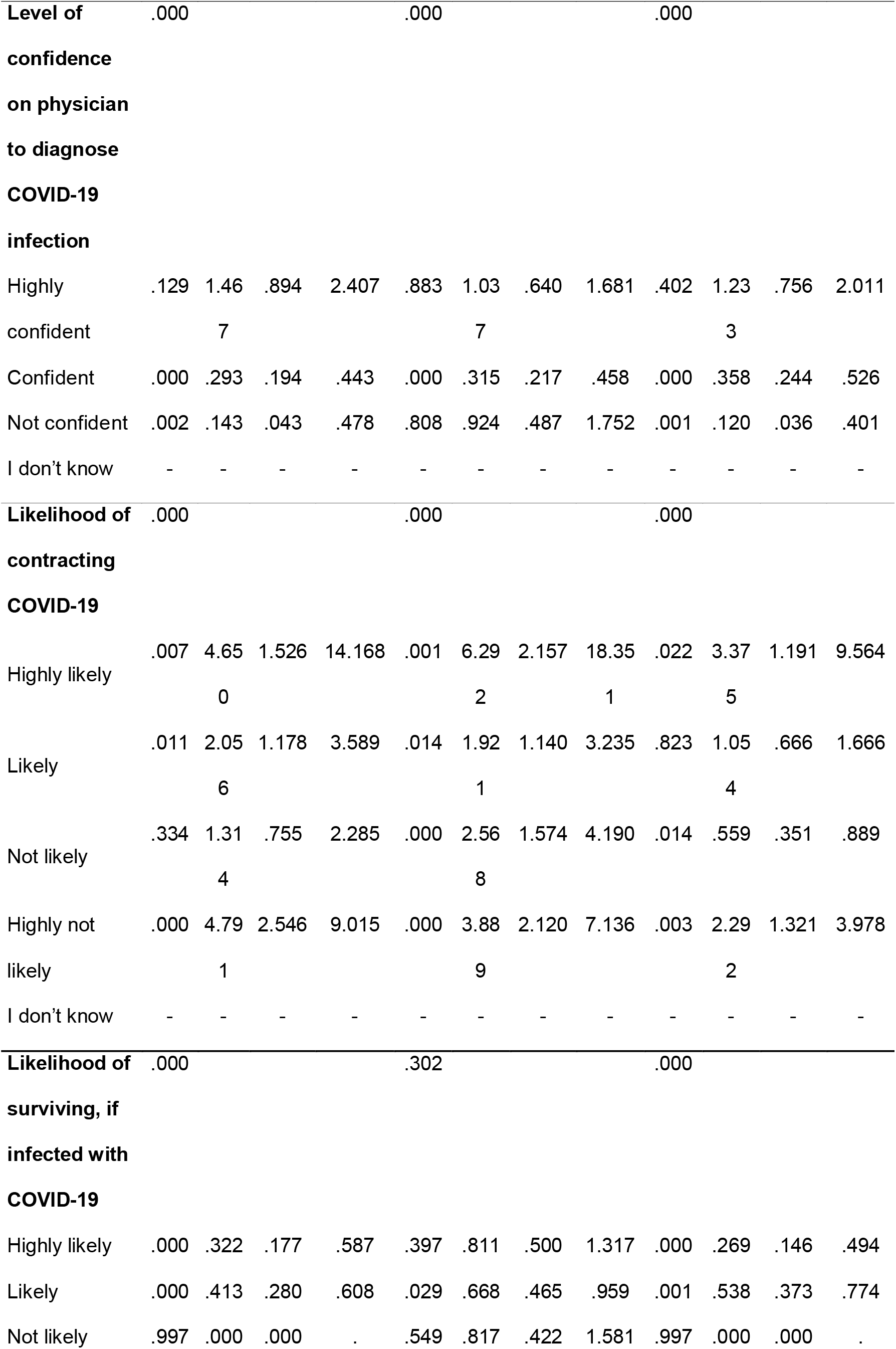
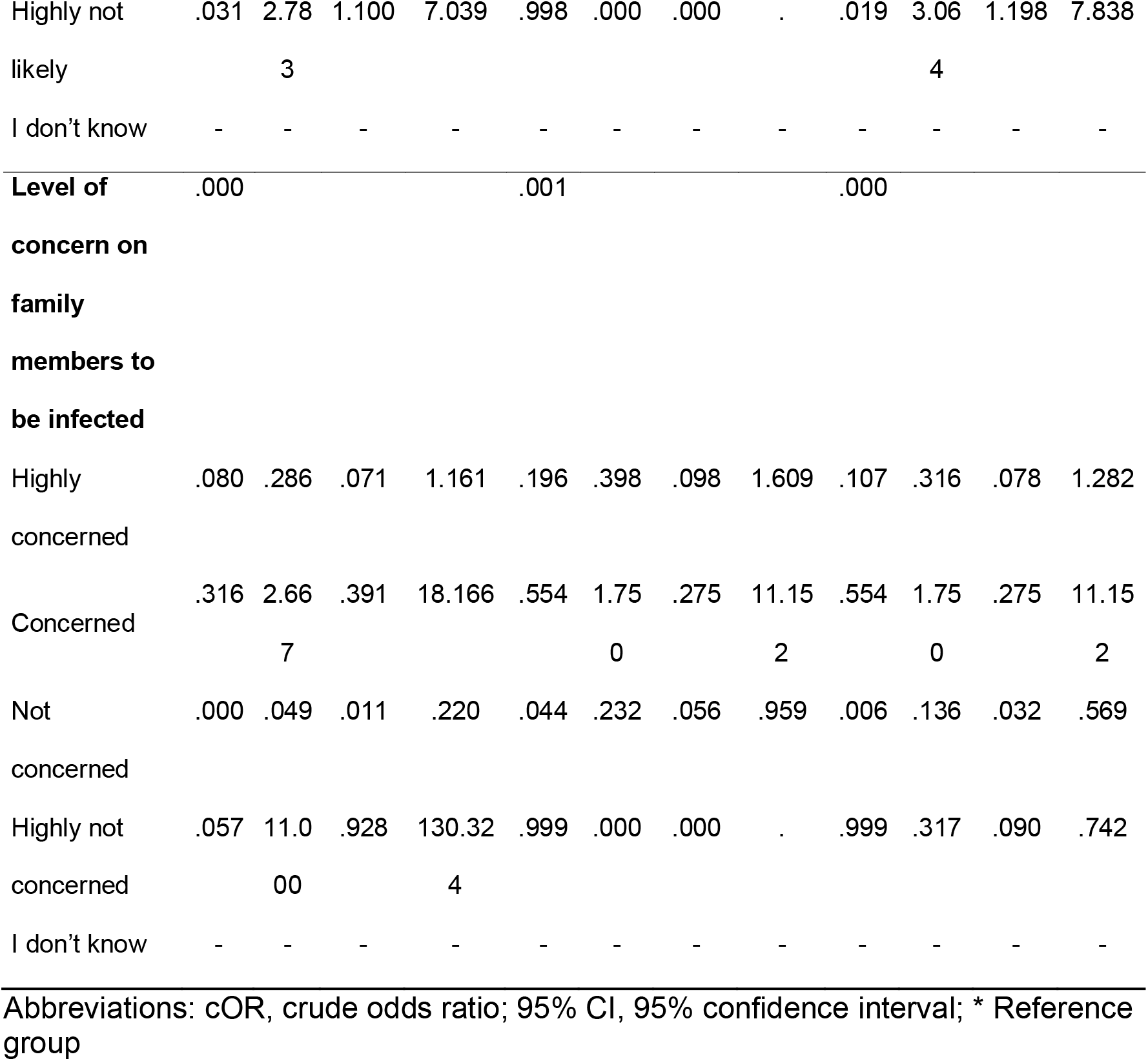
Bivariate analysis of COVID-19 outbreak & lockdown related variables using binary logistic regression

Multiple logistic regression model for depression using stepwise forward LR method was statistically significant, χ^2^(52) = 594.77, P<0.000. The model explained 80.1% (Nagelkerke R^2^) of the variance in depression and correctly classified 94.6% of cases. Six variables were found to be significant predictors including female gender, parental status, frequency of hand washing, level of satisfaction with the information received daily, level of confidence on the physician and likelihood of contracting COVID-19 (Table 4).

**Table 4:**
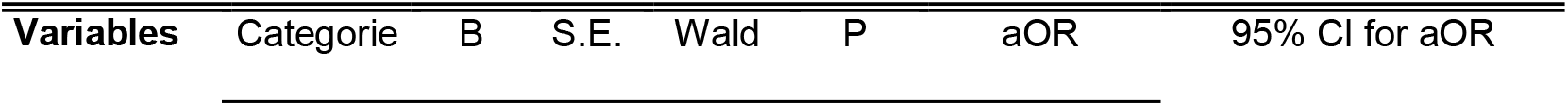
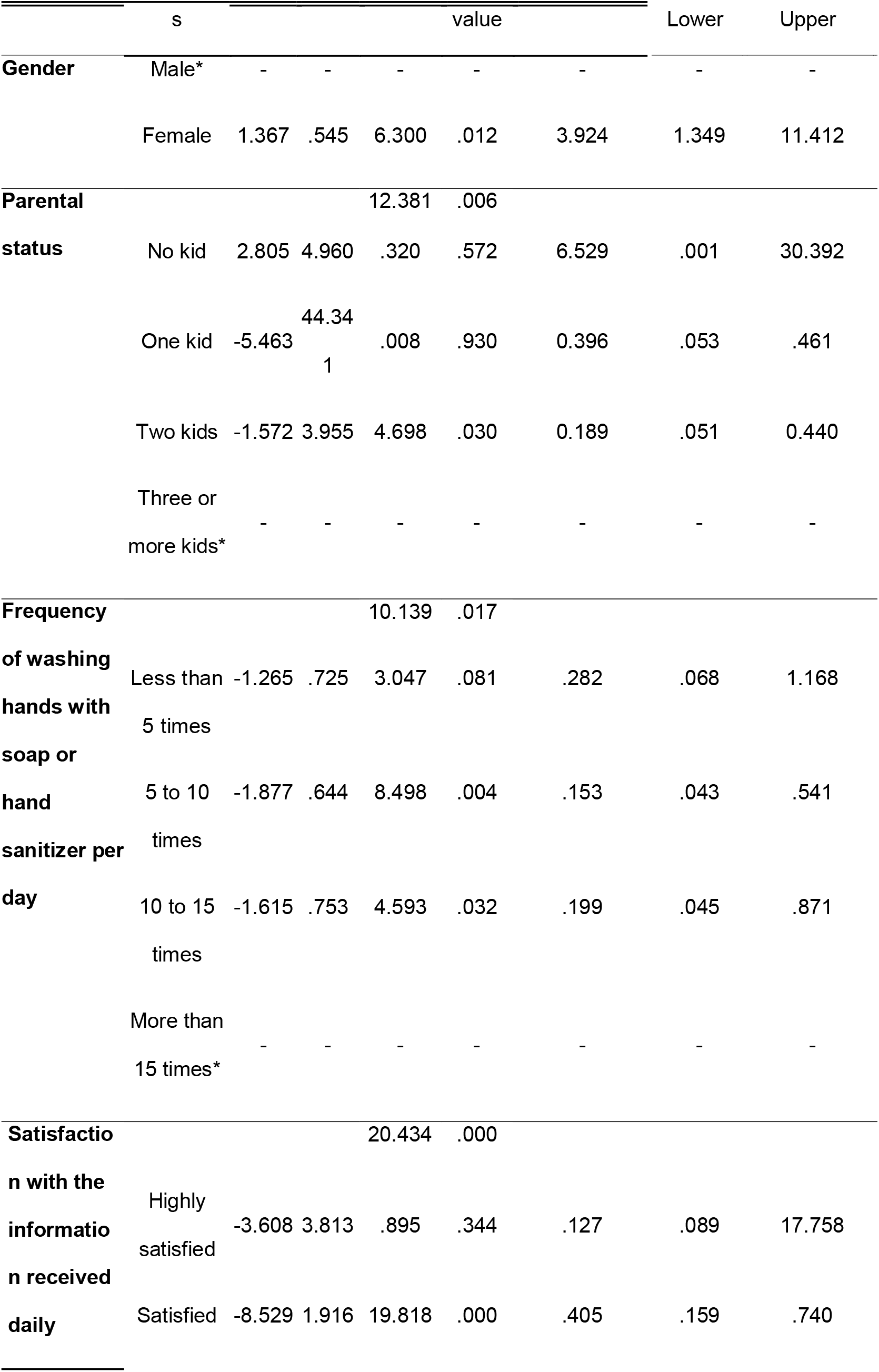
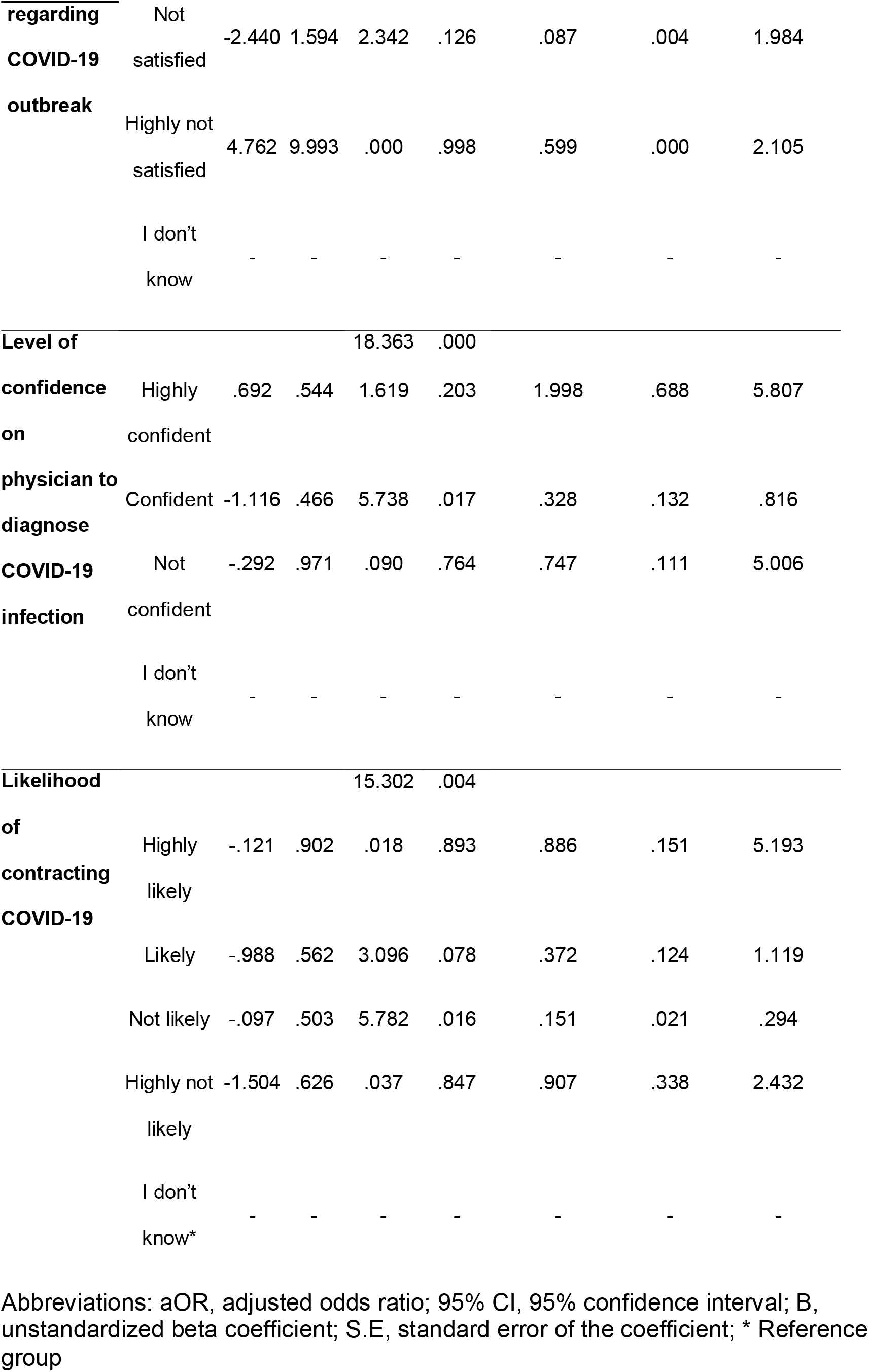
Multiple logistic regression analysis for depression and its correlates

According to bivariate logistic regression analysis, among the sociodemographic variables, marital status (Table 2) and among COVID-19 outbreak & lockdown related factors, wearing masks and gloves in public places, awareness of increase in number of COVID-19 cases, Likelihood of surviving, if infected with COVID-19 (Table 3) were excluded from the final model for anxiety.

The multiple logistic regression model for anxiety using stepwise forward LR method showed significant goodness of fit to our observed data, χ^2^(48) = 455.7, P<0.000. The model explained 59.8% (Nagelkerke R^2^) of the variance in anxiety and correctly classified 86.8% of cases. The significant predictors for anxiety were found to be female gender, educational status, employment status, frequency of checking for information regarding COVID-19, level of confidence on the physician and likelihood of contracting COVID-19 (Table 5).

**Table 5:**
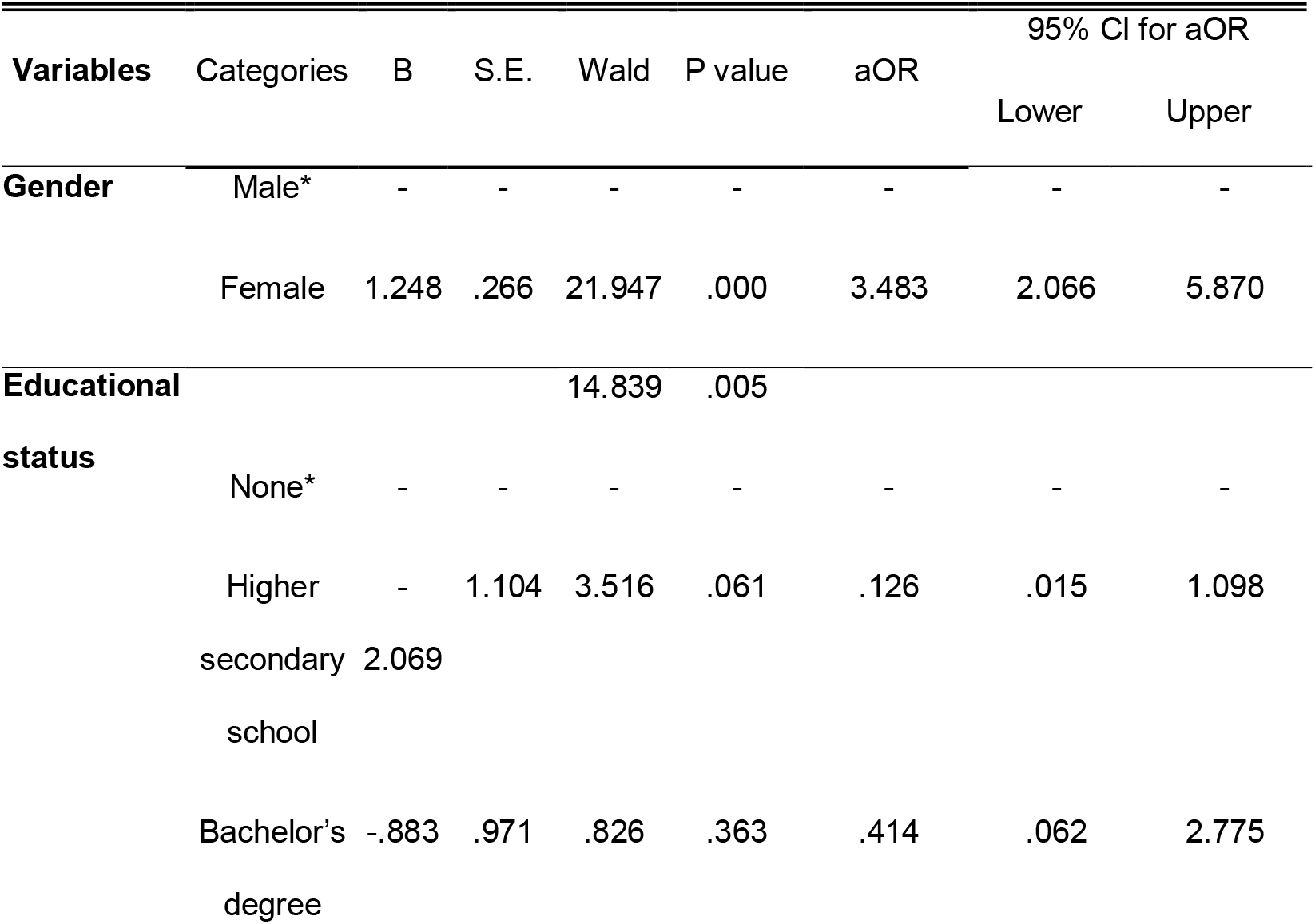
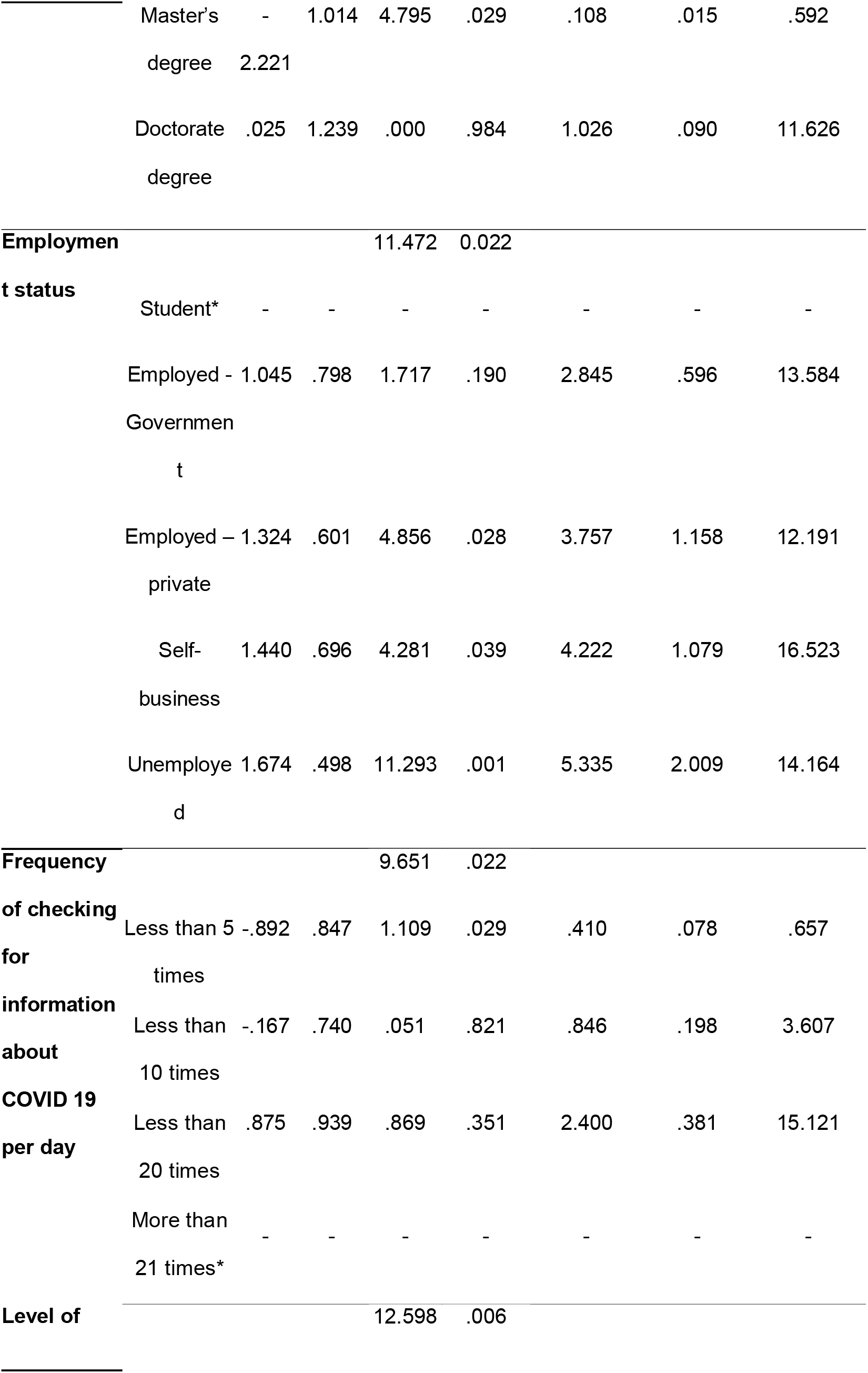
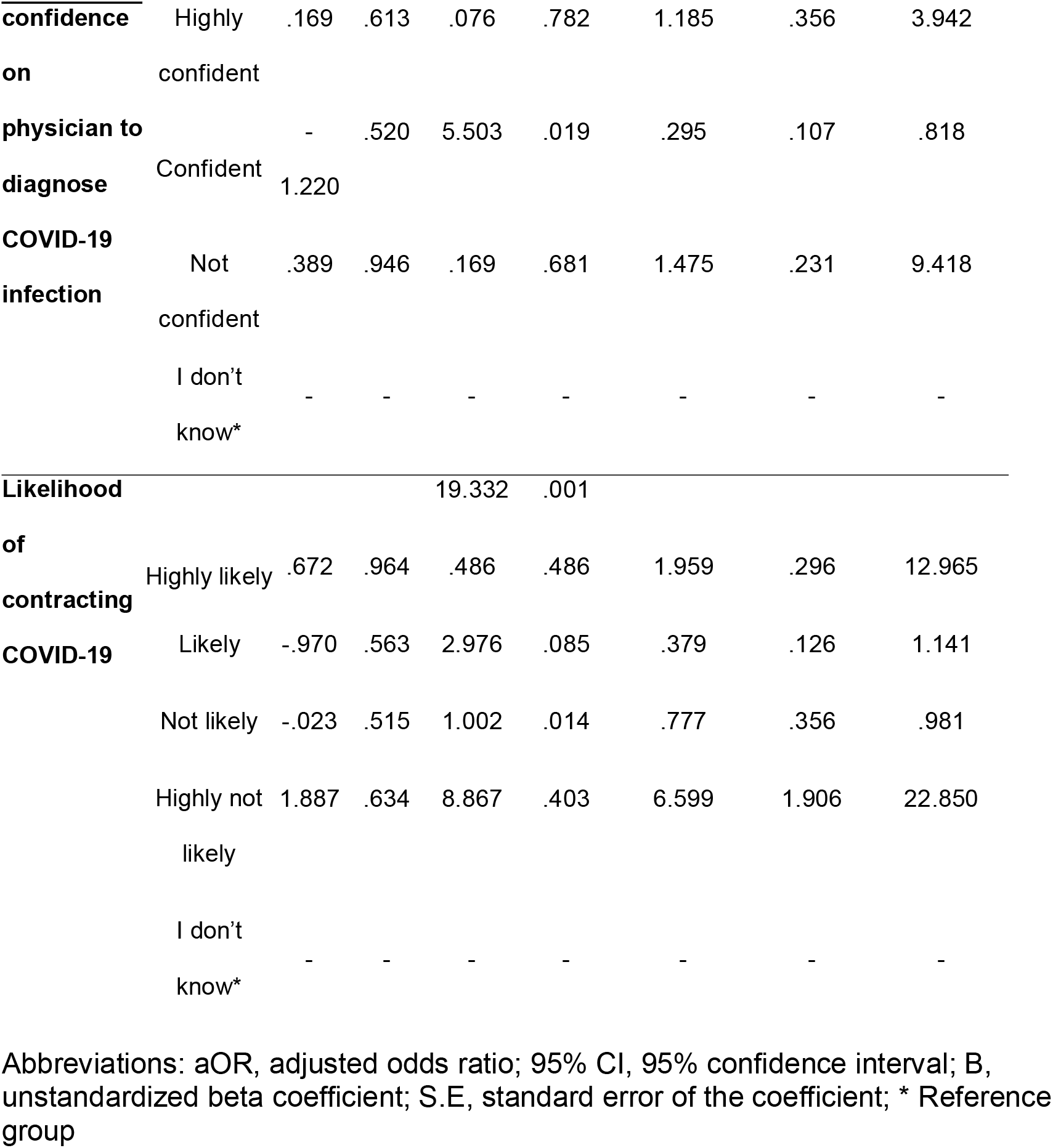
Multiple logistic regression analysis for anxiety and its correlates

Wearing masks and gloves in public places, awareness of increase in number of COVID-19 cases were excluded from the final model for stress based on bivariate logistic regression analysis (Table 3). The multiple logistic regression model for stress using stepwise forward LR method showed significant goodness of fit to our observed data, χ^2^(54) = 621.77, P<0.000. The model explained 78.2% (Nagelkerke R^2^) of the variance in stress and correctly classified 92.1% of cases. Five variables were found to be significant predictors including age of the participants, marital status, household size, level of confidence on the physician and likelihood of contracting COVID-19 (Table 6).

**Table 6:**
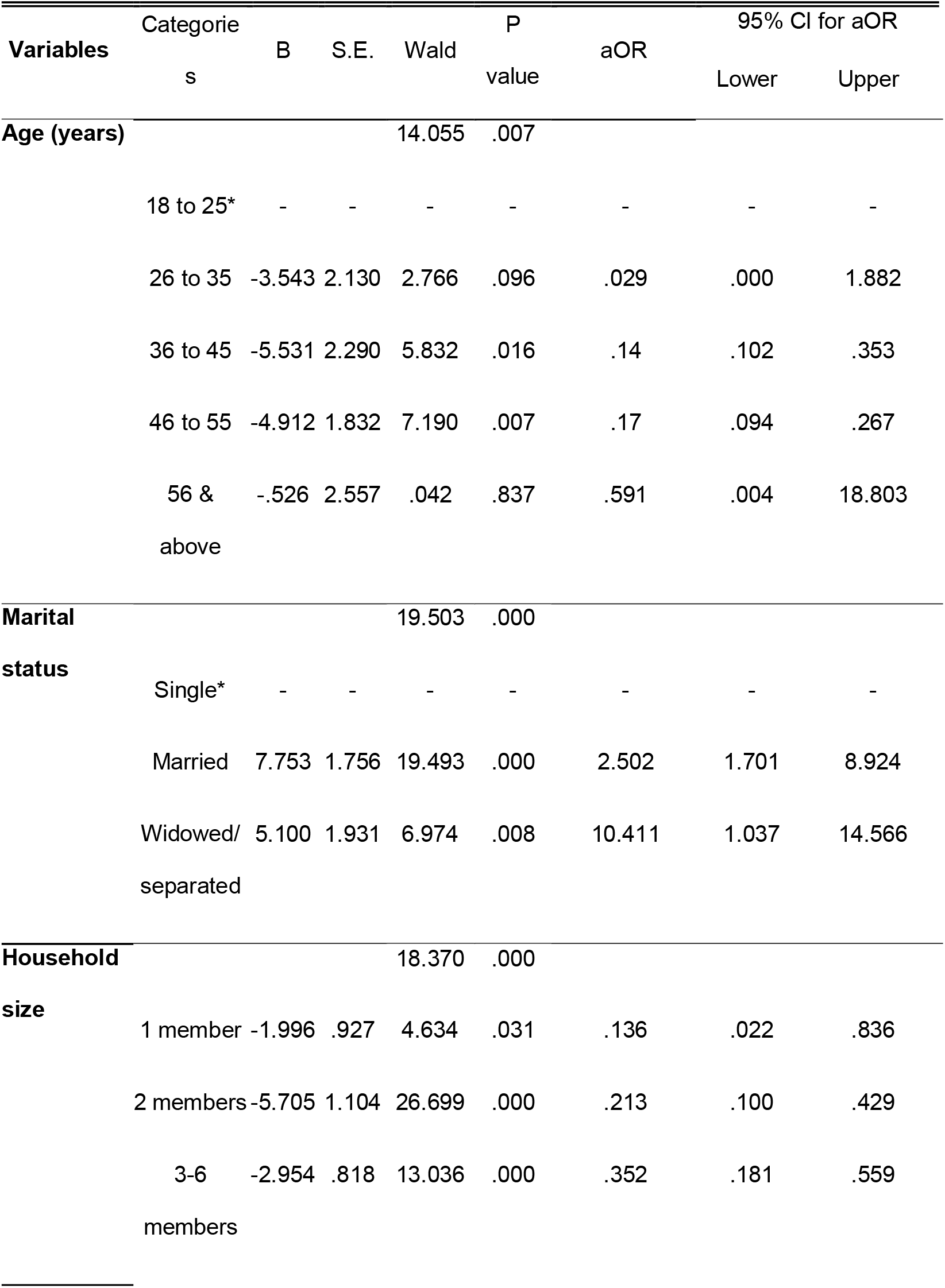
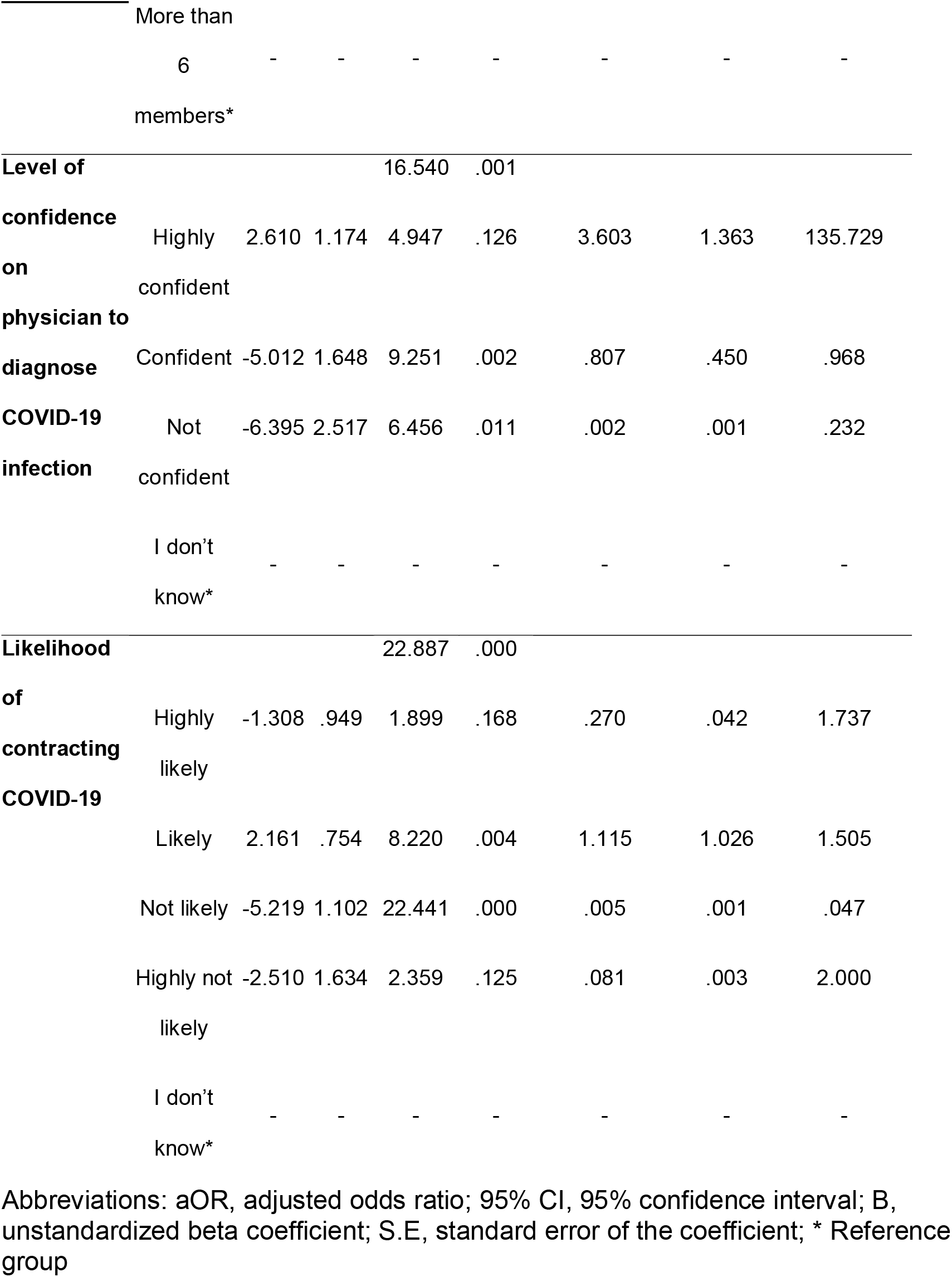
Multiple logistic regression analysis for stress and its correlates

We found that, females were 1.7 times more likely to be present with depressive symptoms (cOR-1.667), 2.6 times as likely to show symptoms of anxiety (cOR-2.624) and twice as likely to show stress symptoms (cOR-2.075) when compared to males but when adjusted for other confounding factors, females were quadruple times as likely as males to have depression (aOR-3.924) and 3.5 times as likely as males to have anxiety (aOR-3.483). Our results are similar to previous studies conducted during SARS and COVID-19 pandemic in Italy and China, where the prevalence of psychological disorders were steadily associated with the female gender [4, 6, 8]. This trend was noted in India before the outbreak too where the prevalence in depression and anxiety disorders were more in females than males [18]. There was no significant association between gender and likelihood of stress which is in accordance with studies conducted during earlier epidemic/pandemic situations [5, 19]. Gender specific help and counselling should be offered to mitigate the psychological strain in the population.

An interesting finding in our study was the variation in association between age and depression, anxiety and stress. Age of the participants was significantly associated with all three domains, where adult Indians of age 36 to 45 years were 1.5 times more likely to have depression (cOR-1.509) and individuals of 46 to 55 years of age were 2.5 times as likely to have anxiety (cOR-2.503) when compared to young adults of age 18 to 25 years. There was also an increase in likelihood of stress with increase in age (36 to 45 years-cOR-1.708, 46 to 55 years-cOR-5.524) with slight decrease in higher risk of stress in age above 55 years (cOR-2.599). However, when controlled for other factors, individuals above 25 years of age were found to be less likely to exhibit symptoms of stress when compared to younger adults of age below 25 years (Table 6]. This finding is similar to studies conducted in similar situations where younger age was found to associated with increased likelihood of stress and psychological distress[5, 6]. Young adults may have trouble coping with drastic societal changes and are more active in social media which is swarming with rumours, which may induce fear, anxiety and other psychological effects. Special focus in identifying and providing mental health help for this age group is imperative.

With increase in level of educational status there was lower likelihood of anxiety but not depression and stress. Those with higher educational qualification were found to be less likely to show anxiety symptoms when compared to those who had none (higher secondary school - cOR-0.337, master’s degree - aOR-0.108). Our finding is similar to the studies conducted both before and in relation to pandemic/epidemic [4, 8, 18, 20]. Educational status influences the occupation and income of the individual, which are in turn associated with psychosocial wellbeing. The economic crisis due to lockdown, might put the individuals without formal education at higher risk of developing anxiety. Counselling, guidance or any form of mental health help should include verbal or pictorial representations to aid this group of population.

Marital status was significantly associated with stress and not with depression and anxiety. Married individuals were 2.5 times as likely (aOR-2.502) and widowed/separated individuals were 10 times as likely as single ones (aOR-10.411) to have stress symptoms. This is in contrary to the studies conducted in China, Iran and Italy during COVID-19 pandemic, where there was no association between marital status and negative mental status[6-8]. However, in India, being married was found to be strongly associated (6 times as likely as single individuals) with increase in the prevalence of mental health disorders in women unlike other neighbouring countries like China and other high resourced countries [21]. Negative psychological effects were observed in separated and widowed individuals [22]. This trend seems to be continuing during COVID-19 quarantine as well which is evident from our results. Married, separated/widowed individuals should be given additional help in combating psychological distress during this pandemic.

Employment status was significantly associated with depression, anxiety and stress where being unemployed and employed as self or in private sector were found to have higher risk of negative mental health when compared to student status. When adjusted for other confounders, unemployed individual were 5 times as likely (aOR-5.335) followed by individuals in self business who were 4 times as likely (aOR-4.222) and private sector employees who were 3.75 times (aOR-3.757) as likely to have anxiety symptoms. This is in contrary to the findings by Wang et al., who found that the student population suffered from higher levels of depression, anxiety and stress during COVID-19 pandemic in China[8]. Uncertainties regarding job security especially in self business and private sector along with the financial strain posed by the quarantine could be the cause for high prevalence of anxiety in the said population.

Monthly income was significantly associated with depression, anxiety and stress, where higher income was found to be protective against negative mental health components. Individuals with monthly income above 100,000INR was less likely to have depression (cOR-0.350) and stress (cOR-0.430) when compared to individuals with income less than 10,000INR per month. This is in accordance with previous studies in India where there was an inverse relation between income and common mental disorders [23]. However, when adjusted for the effects of confounders, there was no independent association between monthly income and depression, anxiety and stress. The relative financial stability in the high-income population could be the factor for relatively lesser psychological distress in the said population.

Having two kids posed lesser risk of depression when compared to having 3 or more kids (aOR-0.189), while there was no significant association with anxiety and stress. However, in bivariate analysis, not having kids and having 1 or 2 kids were found to be protective against depression and anxiety and having one kid or none were found to be protective against stress when compared to having 3 or more children (Table 2). There are varied reports regarding association between parental status and psychological distress. Not having kids was associated with depression during COVID-19 pandemic in Italy [6], having three or more kids were associated with lesser risk of psychological distress during equine influenza epidemic in Australia [5] and having kids was not associated with depression during SARS quarantine in Canada [4]. With the kids being home-schooled, burden on the parents have increased and could be a cause for increase in psychological distress which may increase with the number of kids.

Household size of two members was found to be protective against depression and anxiety and household size of 2 members, 3 to 6 members were protective against stress when compared to bigger household size. When controlled for confounders, individuals from smaller family size viz. one member (aOR-0.136), 2 members (aOR-0.213) and 3 to 6 members (aOR-0.352) were found to be less likely to have symptoms of stress when compared to individuals from family size of more than 6 members. This is in contrary to a previous study in China where there was no association between household size and psychological distress [8, 24]. Lack of personal space and higher financial strain in larger families could be the possible reasons behind our results. Household size should be taken into consideration by the mental health professionals while offering guidance and counselling.

In our study, practice of specific precautionary measures like frequency of washing hands was found to be significantly related to depression, anxiety and stress while wearing masks and gloves in public places had no association when not controlled for confounders. In the final regression model however, lesser frequency of washing hands (less than 15 times) was associated with lesser likelihood of anxiety (Table 5). Previously, anxiety was associated with increase in practice of preventive measures during SARS outbreak [25]. In a survey conducted during 2020 March 22^nd^ −24^th^ in India, 75% of the respondents were found to use gloves and sanitizers, whereas in our study, the number of individuals who do not wear masks and gloves in public places was 11.8% which is higher when compared to China (3.2%) [8, 9]. In our study, 28.9%, 22.1% and 32.3% of participants were not aware that droplets, contact with infected persons and contact with contaminated objects are possible routes of transmission of COVID-19 respectively (Table 1). This lack of awareness could be related to the laxity in practicing personal precautionary measures.

Those who checked for information less frequently (<5 times/day) about COVID-19 were found to be less likely to show symptoms of anxiety when compared to those who checked for more than 21 times a day (aOR-0.410). 74% of the respondents gathered information about the pandemic through internet including social media which gives many inconsistent and fake news which may cause fear and anxiety. Individuals who were satisfied with the information received were found to be less likely to show depression symptoms (aOR-0.405) which was similar to the findings in China [8]. The population should be urged to follow authentic news provided by reliable sources to avoid psychological distress.

Those who felt confident on the physician’s ability to diagnose COVID-19 infection were found to be less likely to have symptoms of depression (aOR-0.328), anxiety (aOR-0.295) and stress (aOR-0.807), similar to the study in China [24]. The confidence in physician’s ability gives a sense of security to the individuals and hence reduces fear and psychological distress.

Individuals who expressed that they were not likely to contract COVID-19 during this outbreak were less likely to have symptoms of depression (aOR-0.151), anxiety (aOR-0.777) and those who thought they were likely to contract COVID-19 were more likely to have symptoms of stress (aOR-1.115). Similarly, the participants who thought that it was highly not likely for them to survive if infected were found to be more likely to have depression (cOR-2.783) and stress symptoms (cOR-3.064) while those who thought it was likely for them to survive had lower levels of anxiety (cOR-0.668) and stress (cOR-0.538). We also found that individuals who were not concerned about their family members contracting COVID-19 were less likely to show symptoms of depression (cOR-0.049), anxiety (cOR-0.232, 95% CI 0.056-0.959) and stress (cOR-0.136). Our findings are similar to previous studies where low levels of perceived likelihood of contracting COVID-19 and surviving the pandemic if infected were protective against depression, anxiety and stress [24]. These concerns should be considered by the mental health workers when providing mental health help for the population.

### Limitations

Our study is limited by the cross-sectional nature and the non-availability of control group. The longitudinal effects of the pandemic and lockdown is not ascertained. The study participants included only those who had access to internet and those who could respond in English. None of our participants were tested positive for COVID-19 and neither had any contact history with known COVID-19 patients. Thus, our findings may not be generalised to the COVID-19 infected patients and their peers.

## Conclusion

To our knowledge, our study is the first to assess the mental health status of the adult Indian population during COVID-19 outbreak & lockdown along with identifying the possible risk and protective factors. During the third phase of the lock down, less than one fifth of the adult Indian population suffered from depression, one fourth suffered from anxiety and more than one fifth suffered from stress. Females were more likely to suffer from depression and anxiety when compared to males. Employment in the government sector and higher educational status were protective against anxiety. Age above 25 years, smaller household size and single status were associated with decrease levels of stress. Parents with lesser number of kids (≤2) or none were less likely to suffer from depression when compared to parents with more than 2 kids. Increased levels of confidence in physician’s ability to diagnose COVID-19 infection, decreased self-perceived likelihood of contracting the infection were associated with decreased levels of depression, anxiety and stress. Less frequency of checking for information on COVID-19 was associated with decreased levels of anxiety and satisfaction of information received about COVID-19 pandemic was associated with decreased levels of depression.

The ripple caused by the COVID-19 outbreak & lockdown will be continuing far into the future and providing mental health support to the population, targeting the vulnerable groups is crucial. Our study provides an expansive assessment of risk and protective factors affecting the mental health of the population, which would help to design strategies and interventional methods to address and mitigate the negative impact of COVID-19 outbreak & lockdown on the mental health of the population and help prevent the same.

## Data Availability

The data is confidential

## Acknowledgement

The authors would like to thank all the participants in this study. The authors would like to thank the Deanship of Scientific Research, Majmaah University for the support of this project.

